# A HYBRID KNOWLEDGE-BASED AND MODIFIED REGRESSION ANALYSIS APPROACH FOR COVID-19 TRACKING IN USA

**DOI:** 10.1101/2020.07.26.20162347

**Authors:** Rafaat Hussein

**Affiliations:** SUNY, Syracuse, NY USA

**Keywords:** covid-19, coronavirus, epidemic, modelling, outbreak, pandemic, simulation

## Abstract

Since its appearance in 2019, the covid-19 virus deluged the world with unprecedented data in short time. Despite the countless worldwide pertinent studies and advanced technologies, the spread is neither contained nor defeated. In fact, there is a record surge in the number of confirmed new cases since July 2020. This article presents a new predictive Knowledge-based (KB) toolkit named CORVITT (Corona Virus Tracking Toolkit) and modified linear regression model. This hybrid approach uses the confirmed new cases and demographic data, implemented. CORVITT is not an epidemiological model, in the sense that it does not model disease transmission, nor does it use underlying epidemiological parameters like the reproductive rate. It forecasts the spread in order to assist the official to make proactive intervention.

## Background

The unfolding covid-19 has turned the world upside down [1] and this unprecedented trend is set to be the worst pandemic of a generation in terms of the increasing number of infected people. In its report on April 7, 2020, the US’ Centers for Disease Control and Prevention (CDC) [2] indicated that the covid-19 poses a severe threat to public health. In its report, the CDC indicated that the “complete clinical picture with regard to covid-19 is not fully known.” To deal with this blurred picture, the World Health Organization (WHO) has compiled an overwhelmingly pertinent database [3]. The CDC provides a daily report that includes new data reported to the CDC by 55 USA jurisdictions [6]. Many other organizations have also provided similar resources and statistics including the Chinese Medical Association Publishing House [4] and the European Centre for Disease Prevention and Control [5]. All the available approaches suggest that the number of covid-19 new cases plays a key role in mapping its trajectory [7] worldwide.

Covid-19 is an evolving epidemic and its up-and-down spreading is a sign of its elusiveness. As of today (July 25, 2020), the covid-19 is striking back with record blows. In general, the covid-19 issue relates to various facets such as public health and social as well as culture characteristics, and the world seems lacking sound methodologies on how to address it. Using predictive tracking quantitative measures can assist the authority to be proactive rather than reactive, thus better prepared to mitigate potential adverse impacts.

## The Model

The literature seems to suggest using the number of new cases and social distance as the key variables to analyze the covid-19 in various ways. In what follows, we provide a background information about the four main covid-19 modeling techniques: system dynamics, agent-based modelling, discrete event simulation, and hybrid simulation [8]. The system dynamics uses differential equation to model resources, knowledge, people, and money, and the flows between these parameters explain the simulation behavior. The agent-based techniques are stochastic, enabling the variability of human behavior to be incorporated to help understand the likely effectiveness of proposed protective measures. The discrete event technique is also stochastic to model operations over time where entities flow through number of activities. The hybrid simulation combines two or more techniques is used for complex behavior. These techniques focus mainly on diseases’ transmission unfolding phases such as quarantine, lock down, testing, and health care services. Some of these approaches are rooted in the literature since 1777, complex, and cumbersome to implement. Without the adequate specialists in advanced and complex mathematical theories and/or computers, the logical question is thus: how could the authorities ascertain the covid-19 spread in order to make a proactive intervention decisions; e.g. to prepare hospitals and intensive care units, to mitigate the adverse impacts of what may happen in the near future? In search for accurate answer and based on the popular utilization of covid-19 relationship between the number of cases [9] and population per land area, the idea of a new index was conceptualized in this study. It represents the number of reported confirmed new cases per population in the specific region the data was recorded. This new concept harnesses the number of cases and the regional crowdedness of people, which varies in the US from a single digit to multi thousand [2]. The index increases with more cases and with more dense populations (shorter social distancing).

In this study, a combined linear regression analysis and data-fitting model is used. To deal with data fluctuation, this model used a short time span of one month maximum for forecasting, [10, 11, 13, and 14]. The data is obtained from the New York Times Journal database [12]. The journal publishes the daily cases of covid-19 by state and county in the US. The data from eleven states was used: New York State (NYS), Florida (FL), California (CA), Colorado (CO), Illinois(IL), Texas (Tx), Louisiana (LA), Washington (WA), Georgia (GA), New Jersey (NJ), and Michigan (MI). Table 1 the data for selected counties in New York State. We first constructed the slope for the confirmed cases and population in each of the states from March 27 to May 11, 2020 and then used polynomials fitting and linear regression analysis for forecasting. Linear Regression is a direct way to deal with the connection between variables. Table 2 shows a typical output from one of the many regression analysis’ runs followed in this study.

**Table 1.** Selected Macro-Data used.

| date | county | cases | county | cases | county | cases | county | cases | county | cases | county | cases |
| --- | --- | --- | --- | --- | --- | --- | --- | --- | --- | --- | --- | --- |
| 3/12/2020 | New York City | 96 | Nassau | 41 | Orange | 1 | Rockland | 7 | Suffolk | 21 | Westchester | 147 |
| 3/13/2020 | New York City | 155 | Nassau | 51 | Orange | 3 | Rockland | 9 | Suffolk | 29 | Westchester | 157 |
| 3/14/2020 | New York City | 269 | Nassau | 79 | Orange | 6 | Rockland | 12 | Suffolk | 41 | Westchester | 178 |
| 3/15/2020 | New York City | 330 | Nassau | 98 | Orange | 6 | Rockland | 13 | Suffolk | 47 | Westchester | 195 |
| 3/16/2020 | New York City | 464 | Nassau | 109 | Orange | 11 | Rockland | 16 | Suffolk | 63 | Westchester | 219 |
| 3/17/2020 | New York City | 646 | Nassau | 131 | Orange | 15 | Rockland | 22 | Suffolk | 84 | Westchester | 379 |
| 3/18/2020 | New York City | 1344 | Nassau | 183 | Orange | 32 | Rockland | 30 | Suffolk | 116 | Westchester | 537 |
| 3/19/2020 | New York City | 2477 | Nassau | 382 | Orange | 51 | Rockland | 53 | Suffolk | 178 | Westchester | 797 |
| 3/20/2020 | New York City | 4419 | Nassau | 754 | Orange | 84 | Rockland | 101 | Suffolk | 371 | Westchester | 1091 |
| 3/21/2020 | New York City | 6226 | Nassau | 1234 | Orange | 163 | Rockland | 262 | Suffolk | 662 | Westchester | 1387 |
| 3/22/2020 | New York City | 9065 | Nassau | 1900 | Orange | 247 | Rockland | 455 | Suffolk | 1034 | Westchester | 1873 |
| 3/23/2020 | New York City | 12329 | Nassau | 2442 | Orange | 389 | Rockland | 592 | Suffolk | 1458 | Westchester | 2894 |
| 3/24/2020 | New York City | 14943 | Nassau | 2869 | Orange | 498 | Rockland | 671 | Suffolk | 1880 | Westchester | 3891 |
| 3/25/2020 | New York City | 20061 | Nassau | 3285 | Orange | 638 | Rockland | 1069 | Suffolk | 2260 | Westchester | 4691 |
| 3/26/2020 | New York City | 23182 | Nassau | 3914 | Orange | 751 | Rockland | 1197 | Suffolk | 2735 | Westchester | 5944 |
| 3/27/2020 | New York City | 25509 | Nassau | 4657 | Orange | 910 | Rockland | 1457 | Suffolk | 3385 | Westchester | 7187 |
| 3/28/2020 | New York City | 30919 | Nassau | 5537 | Orange | 1101 | Rockland | 1896 | Suffolk | 4138 | Westchester | 7875 |
| 3/29/2020 | New York City | 33983 | Nassau | 6445 | Orange | 1247 | Rockland | 2209 | Suffolk | 5023 | Westchester | 8519 |
| 3/30/2020 | New York City | 38375 | Nassau | 7344 | Orange | 1435 | Rockland | 2511 | Suffolk | 5791 | Westchester | 9326 |
| 3/31/2020 | New York City | 43518 | Nassau | 8544 | Orange | 1560 | Rockland | 2863 | Suffolk | 6713 | Westchester | 9967 |
| 4/1/2020 | New York City | 47914 | Nassau | 9555 | Orange | 1756 | Rockland | 3321 | Suffolk | 7605 | Westchester | 10683 |
| 4/2/2020 | New York City | 52400 | Nassau | 10587 | Orange | 2048 | Rockland | 3751 | Suffolk | 8746 | Westchester | 11566 |
| 4/3/2020 | New York City | 57941 | Nassau | 12024 | Orange | 2397 | Rockland | 4289 | Suffolk | 10154 | Westchester | 12350 |
| 4/4/2020 | New York City | 64274 | Nassau | 13346 | Orange | 2741 | Rockland | 4872 | Suffolk | 12328 | Westchester | 13080 |
| 4/5/2020 | New York City | 68726 | Nassau | 14398 | Orange | 3102 | Rockland | 5362 | Suffolk | 12933 | Westchester | 13722 |
| 4/6/2020 | New York City | 73553 | Nassau | 15616 | Orange | 3397 | Rockland | 5703 | Suffolk | 14473 | Westchester | 14293 |
| 4/7/2020 | New York City | 78498 | Nassau | 16610 | Orange | 3599 | Rockland | 5990 | Suffolk | 15561 | Westchester | 14804 |
| 4/8/2020 | New York City | 83673 | Nassau | 18548 | Orange | 3865 | Rockland | 6413 | Suffolk | 15844 | Westchester | 15887 |
| 4/9/2020 | New York City | 89127 | Nassau | 20140 | Orange | 4090 | Rockland | 6665 | Suffolk | 17413 | Westchester | 17004 |
| 4/10/2020 | New York City | 94702 | Nassau | 21512 | Orange | 4532 | Rockland | 7122 | Suffolk | 18692 | Westchester | 18077 |
| 4/11/2020 | New York City | 100840 | Nassau | 22584 | Orange | 4847 | Rockland | 7477 | Suffolk | 19883 | Westchester | 18729 |
| 4/12/2020 | New York City | 105939 | Nassau | 23553 | Orange | 5027 | Rockland | 7721 | Suffolk | 20816 | Westchester | 19313 |
| 4/13/2020 | New York City | 109706 | Nassau | 24358 | Orange | 5182 | Rockland | 7965 | Suffolk | 21643 | Westchester | 19785 |
| 4/14/2020 | New York City | 113632 | Nassau | 25250 | Orange | 5578 | Rockland | 8335 | Suffolk | 22462 | Westchester | 20191 |
| 4/15/2020 | New York City | 121653 | Nassau | 26715 | Orange | 5716 | Rockland | 8474 | Suffolk | 23278 | Westchester | 20947 |
| 4/16/2020 | New York City | 126623 | Nassau | 27772 | Orange | 5888 | Rockland | 8752 | Suffolk | 24182 | Westchester | 21828 |
| 4/17/2020 | New York City | 131003 | Nassau | 28539 | Orange | 6084 | Rockland | 8987 | Suffolk | 25035 | Westchester | 22476 |
| 4/18/2020 | New York City | 135052 | Nassau | 29180 | Orange | 6281 | Rockland | 9171 | Suffolk | 26143 | Westchester | 23179 |
| 4/19/2020 | New York City | 138370 | Nassau | 30013 | Orange | 6394 | Rockland | 9364 | Suffolk | 26888 | Westchester | 23803 |
| 4/20/2020 | New York City | 140881 | Nassau | 30677 | Orange | 6497 | Rockland | 9457 | Suffolk | 27662 | Westchester | 24306 |
| 4/21/2020 | New York City | 143547 | Nassau | 31079 | Orange | 6576 | Rockland | 9568 | Suffolk | 28154 | Westchester | 24655 |
| 4/22/2020 | New York City | 146787 | Nassau | 31555 | Orange | 6705 | Rockland | 9699 | Suffolk | 28854 | Westchester | 25275 |
| 4/23/2020 | New York City | 150327 | Nassau | 32124 | Orange | 6816 | Rockland | 9828 | Suffolk | 29567 | Westchester | 25959 |
| 4/24/2020 | New York City | 155081 | Nassau | 32765 | Orange | 7170 | Rockland | 10091 | Suffolk | 30606 | Westchester | 26632 |
| 4/25/2020 | New York City | 159851 | Nassau | 33798 | Orange | 7988 | Rockland | 11091 | Suffolk | 31368 | Westchester | 27230 |
| 4/26/2020 | New York City | 163106 | Nassau | 34522 | Orange | 8121 | Rockland | 11256 | Suffolk | 32059 | Westchester | 27664 |
| 4/27/2020 | New York City | 165463 | Nassau | 34865 | Orange | 8253 | Rockland | 11366 | Suffolk | 32470 | Westchester | 28007 |
| 4/28/2020 | New York City | 167487 | Nassau | 35085 | Orange | 8389 | Rockland | 11453 | Suffolk | 32724 | Westchester | 28245 |
| 4/29/2020 | New York City | 170124 | Nassau | 35505 | Orange | 8503 | Rockland | 11586 | Suffolk | 33265 | Westchester | 28625 |
| 4/30/2020 | New York City | 172784 | Nassau | 35854 | Orange | 8665 | Rockland | 11708 | Suffolk | 33664 | Westchester | 28969 |
| 5/1/2020 | New York City | 174931 | Nassau | 36161 | Orange | 8766 | Rockland | 11812 | Suffolk | 34037 | Westchester | 29231 |
| 5/2/2020 | New York City | 177490 | Nassau | 36519 | Orange | 8925 | Rockland | 11945 | Suffolk | 34478 | Westchester | 29626 |
| 5/3/2020 | New York City | 179728 | Nassau | 36780 | Orange | 8982 | Rockland | 12025 | Suffolk | 34855 | Westchester | 29884 |
| 5/4/2020 | New York City | 181034 | Nassau | 36965 | Orange | 9030 | Rockland | 12095 | Suffolk | 35077 | Westchester | 30097 |
| 5/5/2020 | New York City | 182318 | Nassau | 37152 | Orange | 9159 | Rockland | 12144 | Suffolk | 35275 | Westchester | 30239 |
| 5/6/2020 | New York City | 183770 | Nassau | 37350 | Orange | 9215 | Rockland | 12204 | Suffolk | 35543 | Westchester | 30426 |
| 5/7/2020 | New York City | 185653 | Nassau | 37593 | Orange | 9343 | Rockland | 12280 | Suffolk | 35892 | Westchester | 30709 |
| 5/8/2020 | New York City | 187157 | Nassau | 37812 | Orange | 9417 | Rockland | 12349 | Suffolk | 36223 | Westchester | 30904 |
| 5/9/2020 | New York City | 188663 | Nassau | 38028 | Orange | 9501 | Rockland | 12400 | Suffolk | 36461 | Westchester | 31086 |
| 5/10/2020 | New York City | 189656 | Nassau | 38217 | Orange | 9558 | Rockland | 12451 | Suffolk | 36702 | Westchester | 31293 |
| 5/11/2020 | New York City | 190546 | Nassau | 38337 | Orange | 9599 | Rockland | 12484 | Suffolk | 36911 | Westchester | 31383 |
| 5/12/2020 | New York City | 191320 | Nassau | 38434 | Orange | 9647 | Rockland | 12504 | Suffolk | 37062 | Westchester | 31471 |
| 5/13/2020 | New York City | 192394 | Nassau | 38587 | Orange | 9708 | Rockland | 12543 | Suffolk | 37305 | Westchester | 31610 |
| 5/14/2020 | New York City | 193663 | Nassau | 38743 | Orange | 9786 | Rockland | 12596 | Suffolk | 37544 | Westchester | 31791 |
| 5/15/2020 | New York City | 195472 | Nassau | 38864 | Orange | 9840 | Rockland | 12637 | Suffolk | 37719 | Westchester | 31942 |
| 5/16/2020 | New York City | 196481 | Nassau | 39033 | Orange | 9894 | Rockland | 12688 | Suffolk | 37942 | Westchester | 32096 |
| 5/17/2020 | New York City | 197486 | Nassau | 39136 | Orange | 9958 | Rockland | 12758 | Suffolk | 38117 | Westchester | 32223 |
| 5/18/2020 | New York City | 198114 | Nassau | 39225 | Orange | 9980 | Rockland | 12777 | Suffolk | 38224 | Westchester | 32322 |
| 5/19/2020 | New York City | 198710 | Nassau | 39295 | Orange | 10003 | Rockland | 12798 | Suffolk | 38327 | Westchester | 32401 |
| 5/20/2020 | New York City | 199392 | Nassau | 39368 | Orange | 10058 | Rockland | 12831 | Suffolk | 38411 | Westchester | 32516 |
| 5/21/2020 | New York City | 200507 | Nassau | 39487 | Orange | 10107 | Rockland | 12877 | Suffolk | 38553 | Westchester | 32672 |
| 5/22/2020 | New York City | 201298 | Nassau | 39608 | Orange | 10157 | Rockland | 12905 | Suffolk | 38672 | Westchester | 32766 |
| 5/23/2020 | New York City | 202062 | Nassau | 39726 | Orange | 10212 | Rockland | 12934 | Suffolk | 38802 | Westchester | 32880 |
| 5/24/2020 | New York City | 202931 | Nassau | 39837 | Orange | 10240 | Rockland | 12963 | Suffolk | 38964 | Westchester | 32967 |
| 5/25/2020 | New York City | 203569 | Nassau | 39907 | Orange | 10259 | Rockland | 12996 | Suffolk | 39090 | Westchester | 33048 |
| 5/26/2020 | New York City | 204111 | Nassau | 39974 | Orange | 10292 | Rockland | 13019 | Suffolk | 39199 | Westchester | 33106 |
| 5/27/2020 | New York City | 204781 | Nassau | 40034 | Orange | 10307 | Rockland | 13047 | Suffolk | 39258 | Westchester | 33185 |
| 5/28/2020 | New York City | 205854 | Nassau | 40140 | Orange | 10340 | Rockland | 13076 | Suffolk | 39359 | Westchester | 33292 |
| 5/29/2020 | New York City | 206800 | Nassau | 40226 | Orange | 10376 | Rockland | 13100 | Suffolk | 39445 | Westchester | 33348 |
| 5/30/2020 | New York City | 207539 | Nassau | 40307 | Orange | 10404 | Rockland | 13128 | Suffolk | 39532 | Westchester | 33428 |
| 5/31/2020 | New York City | 208085 | Nassau | 40396 | Orange | 10421 | Rockland | 13151 | Suffolk | 39643 | Westchester | 33480 |
| 6/1/2020 | New York City | 208550 | Nassau | 40479 | Orange | 10437 | Rockland | 13185 | Suffolk | 39705 | Westchester | 33551 |
| 6/2/2020 | New York City | 209195 | Nassau | 40572 | Orange | 10464 | Rockland | 13223 | Suffolk | 39980 | Westchester | 33632 |
| 6/3/2020 | New York City | 209688 | Nassau | 40644 | Orange | 10475 | Rockland | 13259 | Suffolk | 40062 | Westchester | 33690 |
| 6/4/2020 | New York City | 210227 | Nassau | 40713 | Orange | 10486 | Rockland | 13280 | Suffolk | 40153 | Westchester | 33766 |

**Table 2.** A Sample Output of the Modified Regression Runs.

| Regression Statistics |  |
| --- | --- |
| Multiple R | 0.992092 |
| R Square | 0.984246 |
| Adjusted R Square | 0.976369 |
| Standard Error | 0.372065 |
| Observations | 4 |

| ANOVA |  |  |  |  |  |
| --- | --- | --- | --- | --- | --- |
|  | df | SS | MS | F | Significance F |
| Regression | 1 | 17.29721 | 17.29721 | 124.9503 | 0.007908 |
| Residual | 2 | 0.276865 | 0.138433 |  |  |
| Total | 3 | 17.57407 |  |  |  |

|  | Coefficients | Standard Error | t Stat | P-value | Lower 95% | Upper 95% | Lower 95.0% | Upper 95.0% |
| --- | --- | --- | --- | --- | --- | --- | --- | --- |
| Intercept | -7476.32 | 669.1315 | -11.1732 | 0.007915 | -10355.4 | -4597.28 | -10355.4 | -4597.28 |
| date | 0.170252 | 0.015231 | 11.17812 | 0.007908 | 0.104719 | 0.235785 | 0.104719 | 0.235785 |

### New Knowledge-Based (KB) Toolkit

To accurately and proactively capture the big picture of covid-19 spread, this study transfers the expertise of problem-solving from humans into a KB toolkit that takes in the same data and yields the same conclusion but faster. This new KB-statistic hybrid approach effectively assists humans in dealing with covid-19 massive daily data in addition to save times which is an essential requirement in dealing with the virus illusiveness. The. study introduces for the first time in this field, to our knowledge, a novel KB toolkit to visualize the data and make it easier to understand and use without either mathematical or computer expertise. The CORVITT is a promising incubator for covid-19 future forecasting platforms. Its architecture blueprint emerges from an open-end modular adaptable structure encompassing a graphical-interface client allowing the users to operate it with no professional skills. This KB technology has been proven in other applications and thus applied in this study for covid-19 [15, 16, and 17]. To the author’s knowledge, the concept of CORVITT has not been attempted to date for covid-19. Figure 1 shows the dashboard of CORVITT and Fig. 2 shows the data used in Fig. 1. Although the amount of collected data is massive, the use of the dashboard is intuitive and user friendly. Again, there is no need for medical, mathematical, or computer skills to use the dashboard and benefit from its applications. This is one of the benefits of bringing an artificial brain to help human brains in dealing with complex challenge at hand such as the covid-19.

**Fig 1.**
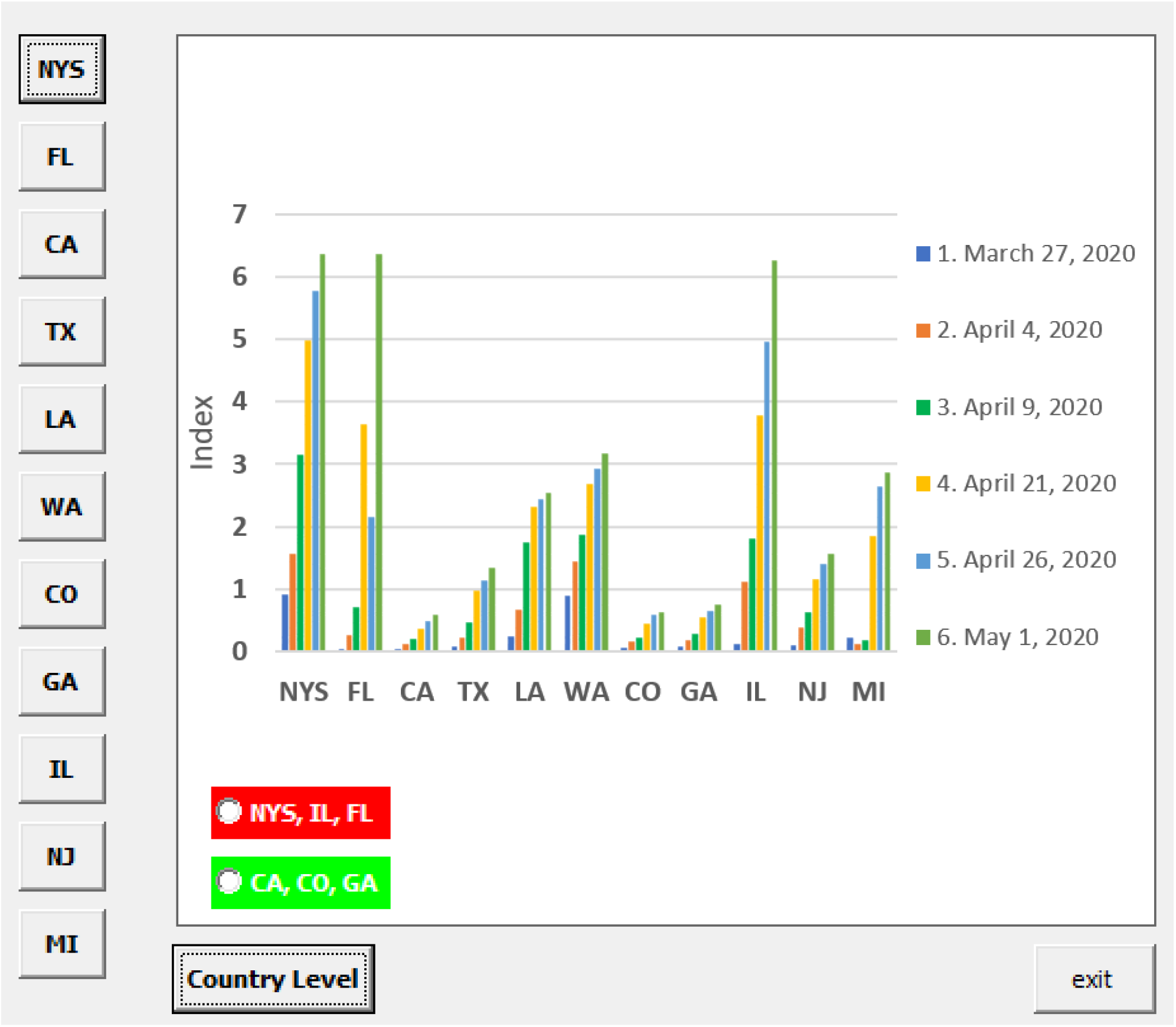
Dashboard of the CORVITT Presented in this Article.

**Fig 2.**
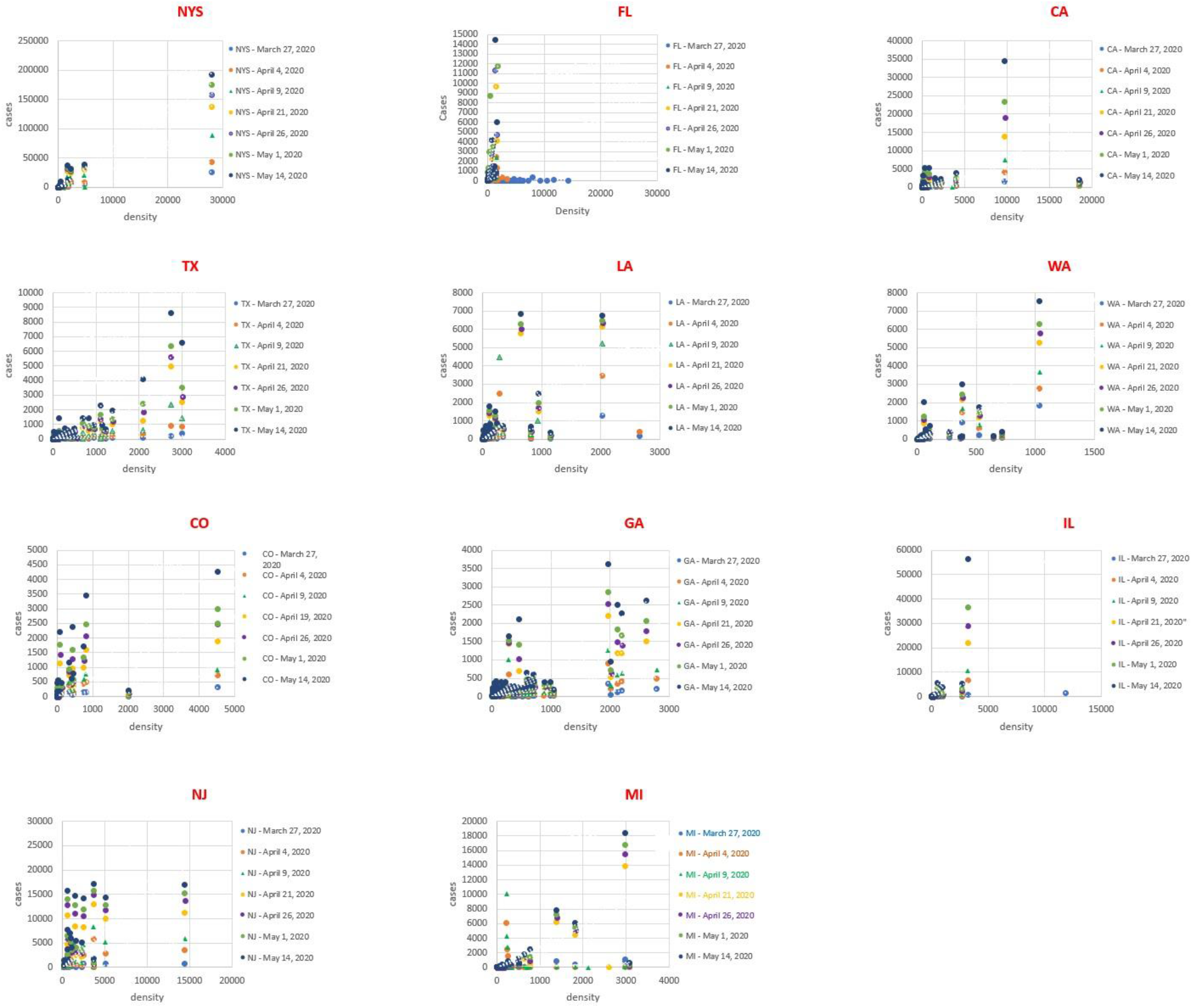
The Macro-Data used in this Study.

## Results and Discussion

In what follows, we examined the feasibility of the ascribed model for covid-19 in two ways: firstly, by analyzing its forecasted outcomes in eleven US states, and secondly by comparing the forecasted results with actual onsite data. Firstly, a database was created at micro-level or counties, for the first time to our knowledge, for the new cases and population per land area on covid-19 from March 27 to May 1, 2020 in the eleven US states: NYS, FL, CA, TX, LA, WA, CO, GA, IL, NJ, and MI. These sates had a steady high number of confirmed cases according to the New York Times Journal. Table 2 shows some of the collected data. Figure 2 shows CORVITT’ gauging of the virus county-wise distributions in terms of the new index and population. Figure 1(d) shows that in NYS, FL, CA, CO, and IL the inhabitants are infected in areas with large social distances because most of the data is concentrated at low population, i.e. large spaces between the inhabitants. On the contrary, in TX, LA, WA, GA, NJ, and MI the virus was spread though the social distance was small because the data spreads over a wide range of population, i.e. large space between the inhabitants. Unlike the common general approach used for all US states at all times since 2019, which is common in the news media from Tabloid to New York Times journals, this discovery unveiled new facets. Figure 1 (d) shows that on March 27, the indexes were 0.93 and 0.10 in NYS and NJ (large distance), respectively, 0.08 and 0.07 for TX and GA (small social distance), respectively, though the spread of data appeared similar on the dashboard in each group. In addition, NYS, LA, WA, IL, and MI have high indexes by comparison to FL, CA, TX, CO, GA, and NJ For example, On April 21, 2020, the indexes were 5.0 and 0.40 for NYS and CA although the closeness of data in both states was similar. Furthermore, the increase of the index within each state was nonuniform. For example, the index increased from 2.8 to 5 between April 9 and 21 in NYS, and from 0.05 to 1.0 in TX over the same period. From a different angel of view, Figure 1(a) shows that the rate of spreading of the virus differ from one state to another. For example, the spreading rate in IL is very high compared to that in CA. This indicates that the virus can affect more people in IL (57920 sq. mi and 13 million people) than NJ (8730 sq. mi and 9 million people). Taken as a whole, CORVITT’ outcomes suggest that NYS is a good region for the virus to spread whereas CA is not as good from March 27 to May 1, 2020. Such information allows the authority to prioritize the resources giving NYS the highest priority. Secondly, Figures 3a and 3b compares the forecasted and actual on-site data and shows a close agreement in different US states. Figure 3a shows that whereas the new cases in NYS has reached the peak in the first week of May, the pandemic was worsening in other states as Illinois, but in California, Georgia, and Colorado reached a plateau. Figure 3c shows the severity of the pandemic as indicated by the skewness; positive (to the right) for LA and negative for NYS, NJ, and MI of the growth and deterioration of the distributions [18]. The positive skewness means longer deterioration (decline in cases) time. The shallow deterioration rate at the trailing end of the curve in Fig. 1(b) is a sign of a plateau. Figure 1 describes the peak, weakness, and steadiness statuses by which the virus trajectory disperses through different stages in various regions. This new discovery is useful to understand the building up and collapse of the virus impacts thus make proactive preparations.

**Figure 3.**
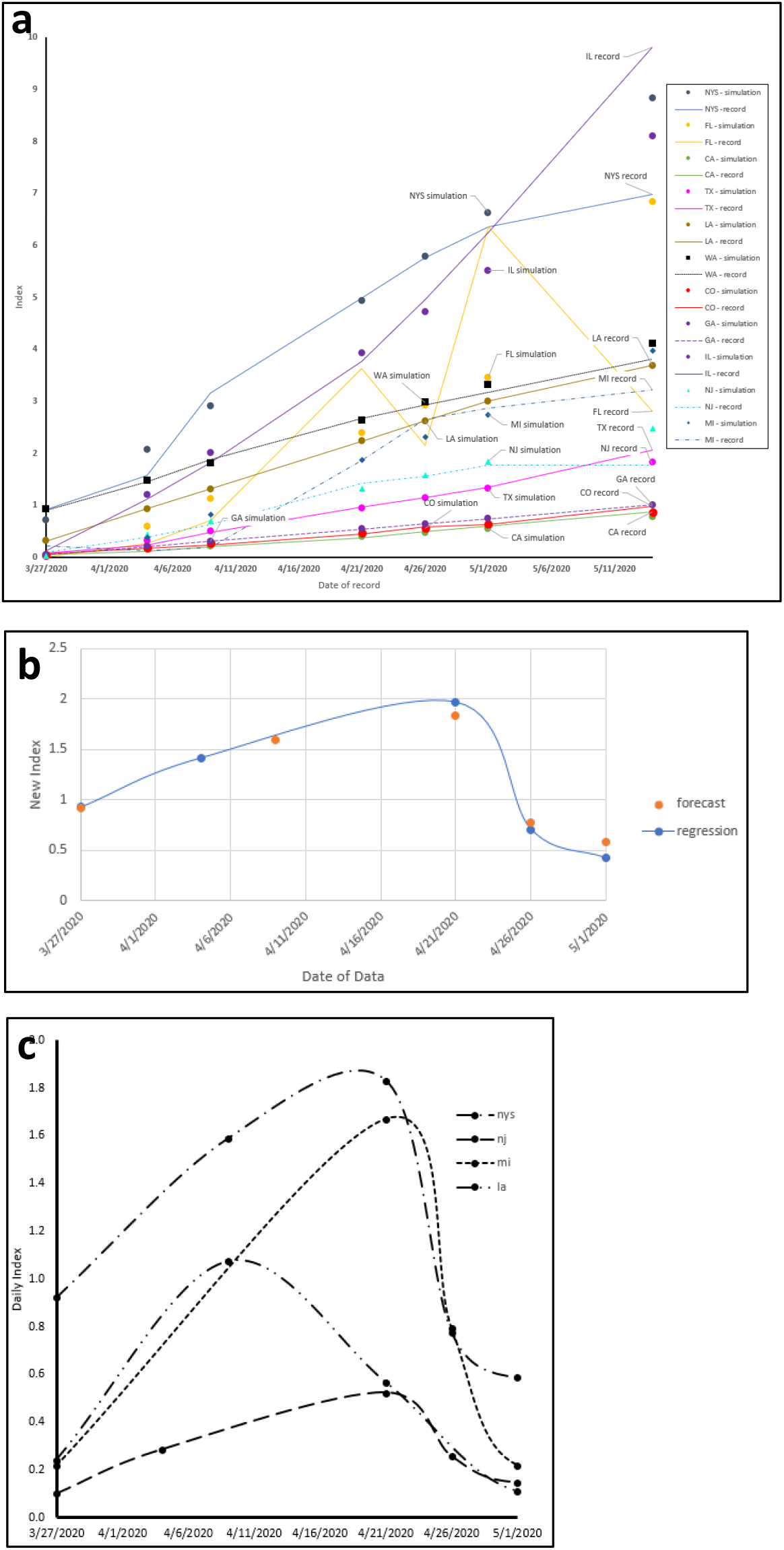
a): The trjectory of cases’ dispersion over time in various US States. b): A satisfactorily agreement between the forecasted and actual data. c). The growth and deterioration distributions of cases over time.

### Summary

As the enormity of the covid-19 threat has become clear, the characteristics of existing covid-19 complex analytic methodologies and the all-encompassing approach place serious limitations on their usefulness for practical use. The computer technologies have reached what no one could imagined, and the KB systems have proven very beneficial in many fields. The logic question is what hasn’t the KB been applied for covid-19 in harness the analytical complexity and easy to apply simulations? This article brought machine smartness to assist humans’ intelligence to capture the big picture of the virus illusiveness thus take proactive steps to mitigate safely its inevitable adverse effects. This article introduced a hybrid KB-regression analysis model for covid-19 forecasting. It used data collected from eleven US states at macro-level level to foresee the short-term spread trajectory. The outputs unveiled new discoveries and shed light on various facets of the covid-19 in each state. The accuracy of the hybrid approach was gauged by comparing forecasted and actual data and satisfactory agreements were found. It should be noted that this study is a step forward, but additional development is in progress for improvement.

### Ethics approval and consent to participate

Not applicable.

### Consent for publication

The author read and approves of the final version of the paper.

### Availability of supporting data

The data used in this study was obtained from the New York Times, https://www.nytimes.com/interactive/2020/us/coronavirus-us-cases.html

### Competing interests

The author declares that he has no competing interests.

### Funding

Not applicable.

### Authors’ contributions

The author wrote the paper and read and approved the final manuscript.

### Author information

Affiliation

State University of New York – ESF, NY

## Data Availability

NA

## Notes

### Competing Interest Statement

The authors have declared no competing interest.

### Clinical Trial

NA

